# Prevalence of gastrointestinal symptoms is higher close to the sea: evidence from coastal Bangladesh

**DOI:** 10.1101/2024.10.18.24315731

**Authors:** Ammatul Fardousi, Masuma Novak, Sharoardy Sagar, Srizan Chowdhury, Rehnuma Haque, Habibur Rahman, Iqbal Kabir, Manzoor Ahmed Hanifi

## Abstract

**Background:** Sea level rise, heavy rainfall, flooding, and temperature changes due to climate change contribute to the spread of enteric infections, disrupting normal intestinal functions and leading to gastrointestinal (GI) symptoms such as nausea, vomiting, and diarrhea etc. Coastal regions of Bangladesh are projected to be highly vulnerable to diarrheal diseases and cholera outbreaks by 2050. However, there is limited research on how coastal proximity affects GI health. Thus, we aimed to examine the association between GI symptoms and geographic distance from the sea in a coastal area of Bangladesh.

**Materials and methods:** This study was conducted in the Chakaria Health and Demographic Surveillance System area of icddr,b which is running since 1999. A total of 61,295 household members were interviewed between 2012 and 2016. GI symptoms were chosen as the primary outcome measure of this study. We employed chi-square tests and logistic regression analysis model.

**Result:** Overall, 7% reported experiencing one or more GI symptoms in the previous two weeks. Diarrhea was the most prevalent symptom (35%), followed by heartburn (22%) and abdominal pain (18%). Prevalence was significantly higher among females (8%, p<0.05), older adults (13%, p<0.05), and individuals from lower socioeconomic backgrounds (8%, p<0.05). Seasonal variation was observed, with the highest prevalence in summer (9%, p<0.05) and the lowest in winter (5%, p<0.05). Additionally, participants living within 15 kilometers (aOR: 1.35, 95% CI: 1.24-1.46) and 15-20 kilometers (aOR: 1.23, 95% CI: 1.10-1.38) of the sea had a higher risk of GI symptoms compared to those residing more than 20 kilometers away, after adjusting for other covariates.

**Conclusion:** Our finding demonstrates that individuals residing near the coast have a higher prevalence of GI symptoms. This evidence suggests the need for targeted public health strategies to improve GI health in climate vulnerable coastal populations. Also, further research is needed to determine the causal effect as well as the underlying biological mechanisms of GI symptoms in these population.

## Introduction

Climate change stands as the paramount global health concern of the twenty-first century, posing significant risks to human health. Bangladesh, a low-lying deltaic nation, is highly vulnerable to the impacts of climate change. The geomorphology of the country, characterized by a vast network of river systems and a coastline of 711 kilometers (km) [1, 2], makes it susceptible to natural hazards [3, 4]. The country’s coastal regions which lie only 1 to 3 meters above mean sea level, are mainly prone to seawater contamination of drinking water sources [3–5]. As sea level rise, water salinity is expected to increase, further exacerbating the risk of high salinity in groundwater [6]. The consumption of saline water has been associated with a range of health effects, including infant mortality, cholera outbreaks, and diarrheal diseases [7, 8].

Climate Change also alters environmental dynamics, such as high ambient temperature, changing precipitation patterns, facilitate the spread and survival of pathogens which directly pose risk to human infectious diseases [9, 10]. Among these, Gastrointestinal (GI) infections account for a large proportion of mortality and morbidity worldwide [11]. Symptoms of GI infections such as diarrhea, vomiting, abdominal pain, and heartburn are prevalent in both developed and developing countries [12–14]. These symptoms ranged from mild to severe, and can lead to shock, coma, and even death particularly to vulnerable populations including children, elderly, and those with weakened immune systems [15].

Diarrheal diseases, in particular, are the second major cause of death among under five children globally, and also the second highest cause of death and disability in low- and middle-income countries [16]. Transmission of diarrheal disease is facilitated by lack of sufficient or safe water, and climate change has the potential to alter the disease frequency and distribution [16]. High temperatures can affect the survival, replication and virulence of pathogen [16]. Additionally, water scarcity, driven by flooding and infrastructure damage, impedes access to safe water, and hygiene practices, further facilitating the transmission of enteric pathogens [17, 18].

By 2050, coastal regions of Bangladesh are projected to be highly vulnerable to diarrheal diseases and cholera outbreaks [19]. These impacts substantially will affect the livelihood and health of people living near to the coast [20]. In Chakaria, tube well water salinity ranges from 34 mg/L to 2000 mg/L, with concentrations exceeding WHO recommended limits and increasing as proximity to the sea decreases [21]. Women residing within 20 (km) of the coastline have a 1.3-fold increased risk of miscarriage [21]. Furthermore, 14% of children aged 6 to 11 months living within 15 (km) of the coast are severely malnourished [21]. Despite these issues, the prevalence of GI symptoms in coastal areas has rarely been examined. Therefore, this study aimed to examine the association between GI symptoms and geographic distance from the sea in a coastal area of Bangladesh.

## Materials and methods

### Settings and study population

This population-based study was conducted in Chakaria upazila (sub-district) in the Cox’s Bazar district, located in the southeastern coastal region of Bangladesh. International Centre for Diarrhoeal Disease Research, Bangladesh (icddr,b) has been operating a Health and Demographic Surveillance System (HDSS) in Chakaria since 1999. The HDSS covers 89,633 people living in 17,955 households across 49 villages. Surveillance worker quarterly visits the households to gather basic demographic information, including birth, death, marriage, and migration [21]. The site is strategically divided into six blocks based on the land category and proximity to the coastline (Fig 1).

**Fig 1.**
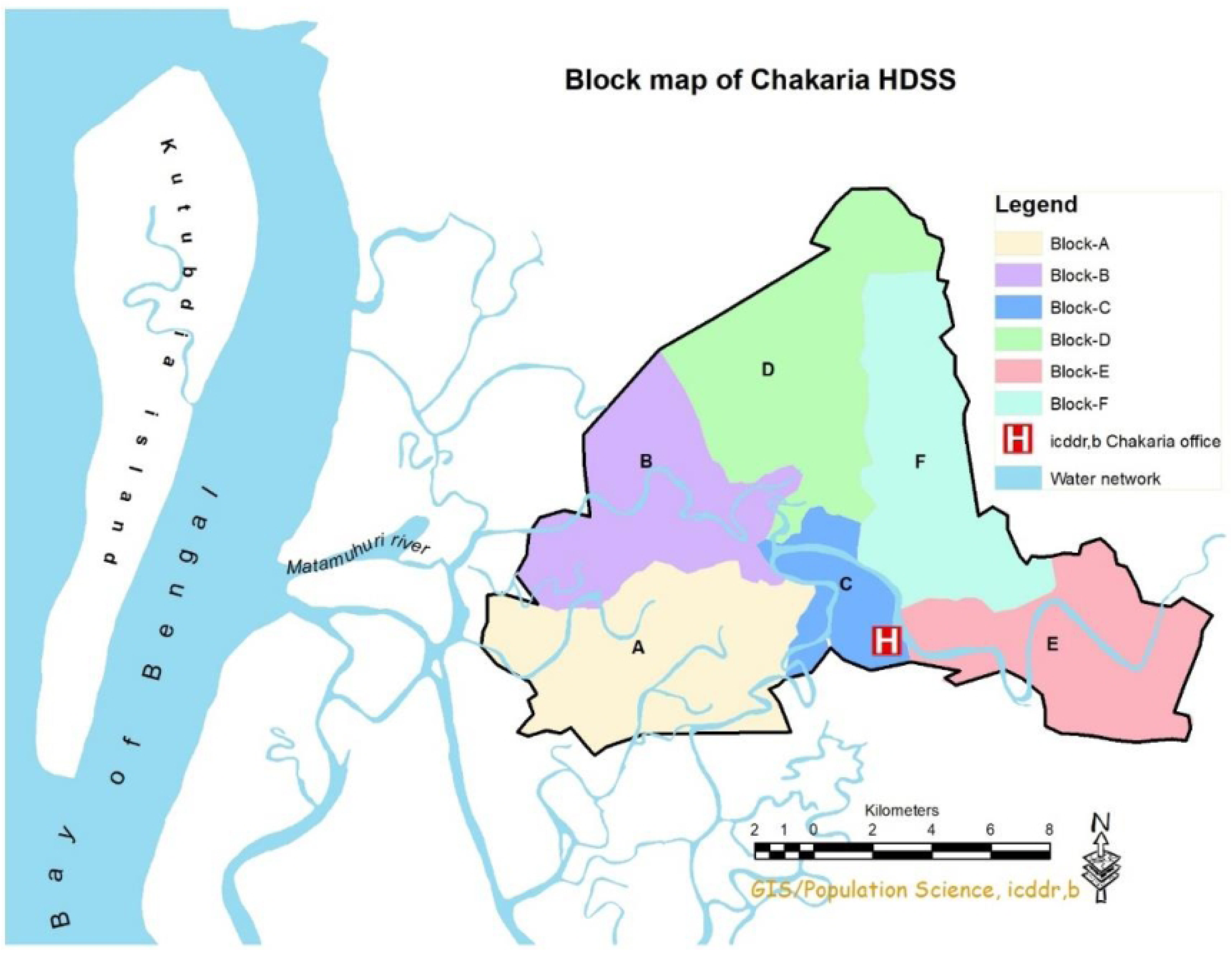
Map of the location of Chakaria Health and Demographic Surveillance System and the Bay of Bengal.

Geographically, Chakaria is similar to other coastal areas in Chittagong district but differs from the rest of the country [22]. It comprises coastal, plain, and hilly areas, and is considered a hotspot to monitor the impacts of climate change on health. Additionally, Chakaria is a relatively low-performing area in terms of health and development indicators compared to central and western parts of the country [22]. In 2011, Chakaria HDSS started collecting geographical information system (GIS) data, recording the geolocations of villages, mosques, temples, hospitals, schools, tube wells, households, rivers, and roads in degree decimal format.

Between 2012 to 2016, we surveyed a total of 61,295 individuals living in 11,398 households in the Chakaria HDSS area. During data collection, respondents were asked whether they had experienced any GI symptoms in the two weeks prior to the interview. Also, information on sociodemographic characteristics were collected. However, asset quintiles and GIS data were retrieved from the existing HDSS database. However, we excluded 1,109 individuals living in 75 households due to the incomplete information of the study variables— i.e., household assets or GIS data. Finally, 60,186 individuals from 11,323 were included in this study (Fig 2).

**Fig 2.**
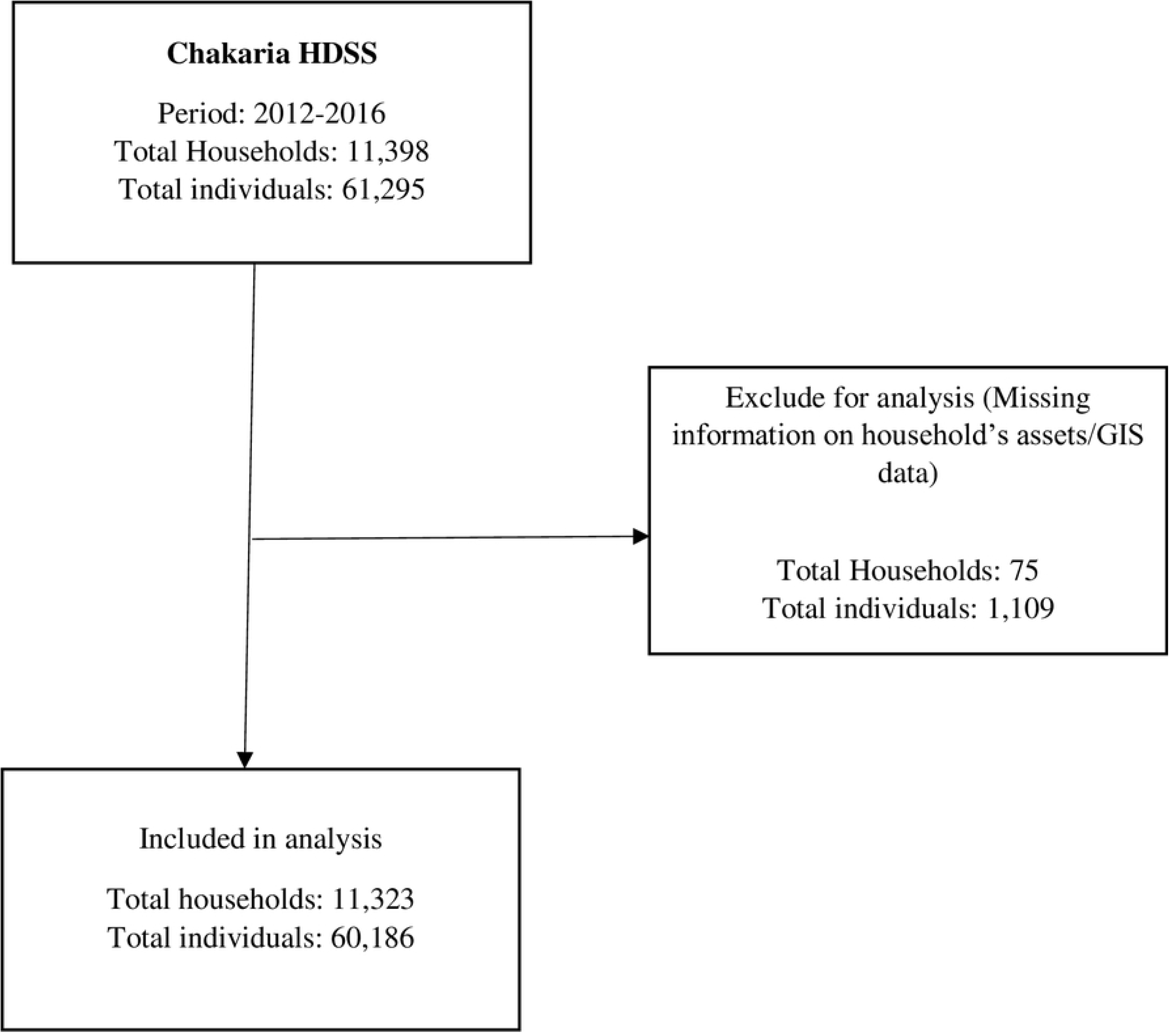
Study population according to inclusion and exclusion criteria (2012-2016)

### Definition of variables

#### GI symptoms

GI symptoms refer to chronic or recurrent complaints affecting different parts of the digestive system, including the pharynx, esophagus, stomach, biliary tract, intestines, or anorectum [14]. In this study GI symptoms were the primary outcome measure, defined as the presence of one or more of the following symptoms e.g., diarrhea, heartburn, abdominal pain, nausea, vomiting, bloating and the passage of mucus with stool.

#### Socioeconomic status (SES)

In this study SES was measured using household asset quintiles, which were determined by the number and types of assets owned by household members. The list includes almirah, table, chair, cycle, bed, motorcycle, electric fan, telephone, television, fridge, sofa, sewing machine, electricity, showcase, and watch or clock.

#### Distance from the sea

Spatial information was collected using GIS to detect and localize the clusters of GI symptoms. To quantify the distance from the sea, the average aerial distance of all households within each village from the nearest shoreline point was calculated and utilized as the metric for the village’s proximity to the sea (Fig 1).

#### Season

We considered these four seasons ‘Summer’ from March-May, ‘Monsoon’ from June-September, ‘Autumn’ from October-November and ‘Winter’ from December-February [23], in accordance with the seasonal patterns recognized in Bangladesh.

### Data analysis

The prevalence of GI symptoms was disaggregated across socio-economic characteristics of the samples, distance from the sea and seasonal variation. The average aerial distance of the villages from the nearest coastline was calculated and categized into 3 groups: <15 km, 15 to 20 km and 20 to 30 km from the sea. Respondent’s age was calculated in completed years from the date of birth. Principal component analysis was used to calculate household asset index scores. All the households were categorized into five equal quintiles, where the first quintile is the poorest 20% of households and the fifth quintile is the wealthiest 20% of households. To see the seasonal variation of the symptom prevalence we categorized it into four groups-summer, monsoon, autumn, and winter.

Chi-square tests were used to investigate the association between the outcome and exposure variables. Multi-level mixed-effect logistic regression was performed to estimate crude odds ratios (cOR) and adjusted odds ratios (aOR) to examine the relationship between geographic distance from the sea and prevalence of GI symptoms, adjusting for age, sex, SES, season, and blocks and villages as a source of random effects. P-values less than 0.05 were considered statistically significant. Frequencies and percentages were used for most variables. To assess the association, we calculated the crude and adjusted odds ratios (OR) with 95% confidence intervals (CI). STATA version 16 was used to analyze data and ArcGIS version 10 was used for creating maps.

### Ethics statement

The studies involving human participants were reviewed and approved by Ethical Review Committee of icddr,b. Informed written consent was obtained from the participants. For the minors, consent was obtained from the parents or guardians.

## Result

### Background characteristics of study population

Of the 60,186 participants included in the study, half of them belong to the age group of 15 to 44 years (45%) and a substantial proportion (12%) of respondents were under 5 years. The proportion of male (49%) and female (51%) was almost equal. After calculating the average distance of all households from the nearest shoreline of the coast, we found that one third of the participants were living within 15 km (27%) from the sea (Table 1).

**Table 1.** Background characteristics of study population, 2012-2016 (N=60,186)

| Characteristics | Proportion (n) |
| --- | --- |
| **Distance from sea (in kilometers)** |  |
| <15 | 26.9 (16,177) |
| 15 – 20 | 39.2 (23,577) |
| 20 – 30 | 33.9 (20,432) |
| **Age (in years)** |  |
| 0 – 4 | 12.4 (7,471) |
| 5 – 14 | 28.1 (16,922) |
| 15-44 | 44.9 (27,027) |
| 45-64 | 10.8 (6,504) |
| 65+ | 3.8 (2,262) |
| **Sex** |  |
| Male | 49.5 (29,779) |
| Female | 50.5 (30,407) |
| **Socio-economic status** |  |
| Lowest | 17.5 (10,526) |
| Second | 18.2 (10,916) |
| Middle | 21.5 (12,949) |
| Fourth | 20.6 (12,412) |
| Highest | 22.2 (13,383) |
| **Season** |  |
| Summer | 40.5 (24,372) |
| Monsoon | 30.2 (18,191) |
| Autumn | 3.1 (1,885) |
| Winter | 26.2 (15,738) |

### Prevalence of GI symptoms

Overall 7% participants reported having experienced one or more GI symptoms in the previous 14 days. Diarrhea (36%), heartburn(21%) and abdominal pain (20%) were the most common symptoms reported. Other symptoms included nausea or vomiting (9%), bloating (9%) and presence of mucus in stool (5%) (Fig 3).

**Fig 3.**
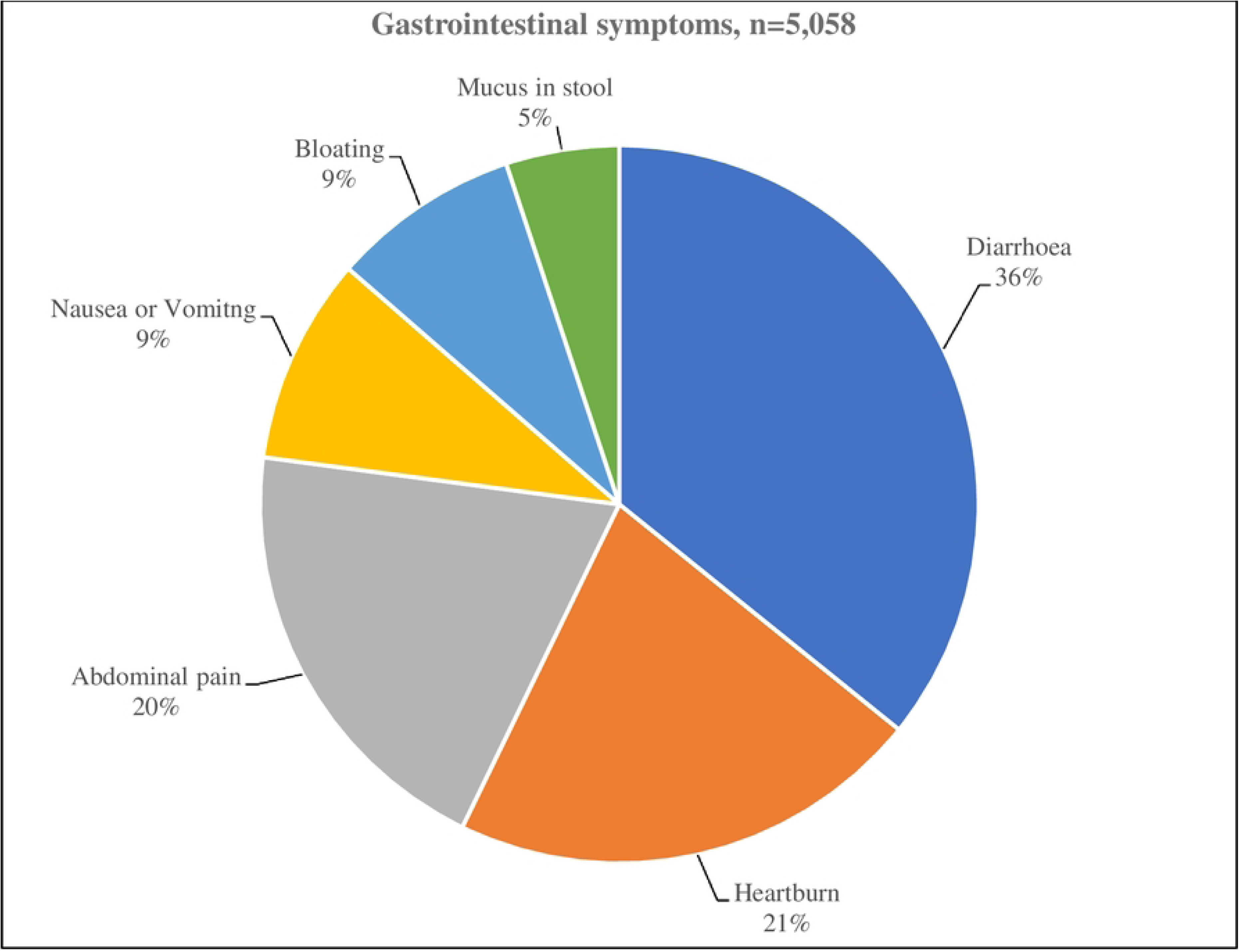
Distribution of Gastrointestinal symptoms in the study population.

### GI symptoms by distance from sea, demographic characteristics, and season

The prevalence of GI symptoms differs significantly according to geographic distance from sea, age, sex, SES and season. Prevalence of GI symptoms was found to decrease with increasing distance from the sea. The highest prevalence observed in participants living within 15 km (9%) from the sea compared to those living more than 20 km (7%) away. The prevalence was higher among females (8%) than men (7%), older people aged ≥65 years (13%), and those from lower socioeconomic background (8%) compared to their other counterparts. The prevalence was highest in summer (9%) and monsoon (8%) compared to winter season (Table 2).

**Table 2.** Percentage of Gastrointestinal symptoms (%) by distance from sea, demographic characteristics, and season.

| Characteristics | % of Gastrointestinal symptoms | P-value |
| --- | --- | --- |
| Distance from sea (in kilometers) |  |  |
| <15 | 8.6 (1,385/16,177) | <0.001 |
| 15 – 20 | 7.8 (1,830/23,577) |  |
| 20 – 30 | 6.6 (1,357/20,432) |  |
| Age (in years) |  |  |
| 0 – 4 | 10.7 (800/7,471) | <0.001 |
| 5 – 14 | 4.3 (731/16,922) |  |
| 15-44 | 7.3 (1,970/27,027) |  |
| 45-64 | 11.8 (768/6,504) |  |
| 65+ | 13.4 (303/2,262) |  |
| Sex |  |  |
| Male | 6.8 (2,036/29,779) | <0.001 |
| Female | 8.3 (2,536/30,407) |  |
| Socio-economic status |  |  |
| Lowest | 8.3 (874/10,526) | <0.001 |
| Second | 8.3 (907/10,916) |  |
| Middle | 7.6 (1028/12,949) |  |
| Fourth | 7.4 (913/12,412) |  |
| Highest | 6.6 (889/13,383) |  |
| Season |  |  |
| Summer | 8.8 (2,140/24,372) | <0.001 |
| Monsoon | 8.3 (1,510/18,191) |  |
| Autumn | 5.9 (112/1,885) |  |
| Winter | 5.2 (810/15,738) |  |

In regression model, we observed that participants residing within 15 km from the sea, there was 35% (95% CI: 1.20-1.51) more risk of suffering from GI symptoms compared to those living between 20-30 km. After adjusted for age, sex, SES and the season the odds for experiencing GI symptoms according to geographic distance from sea were 37% ( 95% CI: 1.20-1.57) higher near to the coast (Table 3).

**Table 3.** Percentage of crude and adjusted* odds ratio according to distance from sea.

| Distance from sea (in kilometers) | Crude odds ratio (95% CI) | Adjusted odds ratio (95% CI) | P-value |
| --- | --- | --- | --- |
| <15 | 1.35 (1.20 - 1.51) | 1.37 (1.20 - 1.57) | <0.001 |
| 15 – 20 | 1.19 (1.07 - 1.31) | 1.23 (1.10 - 1.38) | <0.001 |
| 20 - 30 | Ref | Ref |  |

\*Adjusted for age, sex, SES and season

## Discussion

This is the first population-based study to estimate the association between GI symptoms and geographic proximity to the sea in a coastal area of Bangladesh. We observed that 7% of participants reported experiencing GI symptoms within the previous 14 days. There was a 37% (95% CI, 1.20-1.57) higher risk among those living closer to the sea (within 15 km).

Global studies have reported varying prevalence rates of GI symptoms, ranging from 14% to 61% [24–28]. However, the observed variations in the estimated prevalence across different countries can be attributed to multiple factors including differences in study population, lifestyle, diagnostic criteria and recall period [29]. Moreover the recall period for symptom reporting varied among studies, from one week in USA [24], two weeks in Iran [26] to one month in Turkey [27]. In case of study population, USA [24] and Turkey [27] included individuals above 18 years, in India the focus was on elderly people aged 45 or above [28] and Iran included participants from all age [26]. Our study included all age group population in coastal area and the recall period was 14 days. While another study in Bangladesh by Praveen et al., in individuals aged over 15 years used Rome III diagnostic criteria for adult functional dyspepsia and focused on upper GI symptoms, where the recall period was 3 months [25].

In our study, diarrhea, heartburn, and abdominal pain were the most frequently reported GI symptoms. This finding aligns with Sezgin et al., [27] where the most prevalent GI symptom reported was heartburn (29%). However, abdominal pain is a common complaint across countries [30]. The increased risk of GI symptoms among those living closer to the sea is linked to several factors including limited access to safe water, proper hygiene practices, which facilities the transmission of enteric pathogens [9, 10]. Additionally, a significant association was found between high salinity in drinking water and hospital visits for diarrhea and abdominal pain [3]. Moreover, disruptions in the gut microbiome, caused by environmental stressors such as exposure to contaminated and saline water, are increasingly recognized as a factor in gastrointestinal diseases. These disturbances can lead to dysbiosis, reducing microbial diversity and compromising immune function, thereby heightening susceptibility to GI symptoms [31, 32]. Together, these factors create a complex interplay of environmental and biological risks that disproportionately affect coastal populations, explaining the increased prevalence of gastrointestinal issues in these areas.

In this study, age, sex, SES and season were significantly associated with GI symptoms. The prevalence of GI symptoms was highest in elderly people aged ≥ 65 years. This finding aligns with reports by Chaplin et al., and Zuchelli et al., who also observed a high prevalence of GI symptoms in ageing population [33, 34]. This can be attributed to the ageing-related physiological changes in the GI system, including alterations in motility, releasing enzyme and hormone, digestion, and absorption, as well as pathological condition which are more prevalent in this age group [28, 35].

However, Gl symptoms are gender-specific, and our study observed higher prevalence among women which is consistent with other studies as well [27, 36]. Hormones, particularly estrogen affects the intestinal microbiomes and immune cells, and menstrual cycle affects GI transit duration, leading to GI symptoms [36]. Additionally, Stuart et al., found that low social class is linked with GI symptoms, which aligns with our study findings [37]. Several factors contribute to this disparity, including limited access to clean water, sanitation facilities, and healthcare resources among economically disadvantaged populations [14]. The prevalence of GI symptoms was highest during the summer season in or study. Several factors may contribute to this finding. Rising atmospheric temperature and changing weather patterns in summer exacerbate challenges related to nutrition and clean water [38]. A study conducted by Na et al., found a strong association between temperature and infectious diarrhea [39]. Also, due to the increase in rainfall events in summer, enteric infections rise accordingly [38]. Because these two events eventually alter the survival, replication, virulence and mobilization of enteric pathogen [16].

### Strengths and limitations

Our study utilized data from a coastal HDSS area with a large sample size, which makes the observed relationship more reliable. And we mapped spatial distribution of GI symptoms based on proximity to the sea, providing insights into geographic patterns. And this study is the first to explore this link, providing insights in an under-research area in Bangladesh. There are some shortcomings in our study. Firstly, we relied on self-reported data, which introduces the potential for recall bias and underreporting of GI symptoms. Additionally, our study includes cross sectional study design, which limits the ability to identify the causal relationships between risk factors and GI symptoms.

## Conclusions

We conclude that individuals residing near the coast have a higher prevalence of GI symptoms. This evidence suggests the need for targeted public health strategies to improve GI health in climate vulnerable coastal populations. Also, further research is needed to determine the causal effect as well as the underlying biological mechanisms of GI symptoms in these population.

## Data Availability

The data supporting the conclusions of this article will be made available according to icddr,b’s data access policy.

## Acknowledgments

We would like to thank the villagers and team members of Chakaria HDSS for their cooperation. We gratefully acknowledge icddr,b, as well as the Governments of Bangladesh and Canada for providing core/unrestricted support to icddr,b.

